# Prayer, Politics, and Policy Related to Age-Adjusted Cancer, Heart Disease, Infant Mortality, and COVID-19 Death Rates, U.S. States 2018-2021

**DOI:** 10.1101/2024.10.13.24315408

**Authors:** Leon S. Robertson

## Abstract

The role of religion and politics in the responses to the coronavirus pandemic raises the question of their influence on the risk of other diseases. This study focuses on age-adjusted death rates of cancer, heart disease, and infant mortality per 1000 live births before the pandemic (2018-2019) and COVID-19 in 2020-2021. Eight hypothesized predictors of health effects were considered by examining their correlation to age-adjusted death rates and indicators of health behavior among U.S. states, percentage who pray once or more daily, Republican attitudes and influence on state health policies as indicated by the percentage vote for Trump in 2016, percent of household incomes below poverty, median family income divided by a cost-of-living index, the Gini income inequality index, urban concentration of the population, physicians per capita, and public health expenditures per capita. Since prayer for divine intervention is common to otherwise diverse religious beliefs and practices, the percentage of people claiming to pray daily in each state was used to indicate potential religious influence. Based on collinearity, inequality was chosen for inclusion over poverty, and the prayer and political variables were analyzed separately. All of the death rates were higher in states where more people claimed to pray daily. Only cancer and COVID-19 were correlated significantly with Trump’s percentage of the vote. Lower death rates from cancer and heart disease are associated with more public health expenditures, but not for infant mortality or COVID-19 deaths. COVID-19 death rates were lower in states with more physicians per capita, but that variable was not significantly associated with the other death rates. Heart disease, infant mortality, and COVID-19 death rates were higher in states with more income inequality. All rates except infant mortality were lower in states where a greater percentage of the population resides in urban areas. The correlation between daily prayer and smoking cigarettes, as well as the neglect of public health recommendations for fruit and vegetable consumption and COVID-19 vaccination, suggests that reliance on prayer may be a factor in neglect of preventive practices.

## Introduction

The coronavirus pandemic tested medical, political, and economic resources worldwide. Despite its economic and scientific resources, the U.S. had among the highest death rates in democratic countries. The U.S. Centers for Disease Control and Prevention’s response mainly consisted of recommendations regarding distancing, mask use, testing, and consultations with local health officials [1]. Unreliable testing equipment sent to the States had to be recalled, delaying the testing of the sick [2]. Masking and distancing recommendations were widely resisted, especially where they were mandated [3]. On-demand testing was associated with increased virus spread as negative tests were promoted to make travel decisions, with insufficient emphasis on isolation if one tested positive (4). The public health system, underfinanced in many states, was unprepared to identify and contact persons exposed to those infected. On 70 percent of the days in U.S. states in 2020-2021, there were not enough tracing personnel to interview the infected, much less trace their contacts [5]. People with negative tests traveled and came in contact with infected people who would not isolate or had no symptoms [4]. The major accomplishment of the federal government was the effort to develop vaccines that turned out to be effective beyond expectations. Getting people to use them was more difficult [5,6].

After a CDC spokesperson issued severe warnings about the virus in February 2020, then-President Trump took charge of televised public briefings, claiming that the virus would disappear and endorsing treatments lacking scientific support [7]. Following the initial nationwide shutdown in spring 2020, authority over restrictions on travel, gathering sizes, mask mandates, and vaccination requirements shifted to the state and local governments, as did the procurement of protective gear for healthcare workers [8]. These issues became politicized. Testing and political impacts were separate but additive [9]. When accounting for crowding in various venues and other risk factors, COVID-19 death rates were higher in counties with more negative tests per capita and, independently, in those with a greater percentage of Trump votes [10]. Most Republican governors, with some exceptions, were less likely to impose restrictions, and a few even questioned vaccine effectiveness [11]. Fox News and right-wing radio broadcasts promoted skepticism of science [12]. A poll showed that confidence in the CDC’s accuracy among Fox News viewers dropped from about 85 percent in early 2020 to 19 percent two years later, while remaining above 80 percent among CNN and MSNBC viewers. By 2022, only 16 percent of Fox News viewers reported wearing masks, compared to 60 percent among viewers of other networks [13]. In the Summer of 2021, around forty percent of surveyed healthcare professionals received threats [14]. Restricting attendance at religious services was especially controversial [15], with some religious leaders opposing mask use and vaccines [16].

In recent decades in the U.S., religious fundamentalists have increasingly influenced Republican politics. Republican operatives and a fundamentalist evangelist formed an organization called The Moral Majority that persuaded many evangelistic clergy and their followers to support Republican candidates. However, most African American fundamentalists and some religious liberals remained loyal to the Democratic Party [17]. By the second decade of the 21st Century, Republicans had about a 20-percentage-point advantage among Protestants [18,19].

The scientific study of religious beliefs and practices regarding diseases and injuries mainly focuses on whether individual risk is associated with prayer or attending religious gatherings. The results are mixed. A large-sample longitudinal study of cardiovascular disease among women found that, adjusted for other risk factors, prayer and reading religious literature were associated with subsequent increased cardiovascular risk [20]. A study of African-American church attendance and subsequent mortality found that deaths were lower among regular attendees [21]. Church attendees were found to live longer in another study, but shorter lives were associated with private religious practices [22]. Deaths per population less than 75 years of age were substantially correlated with the percentage of people who say they pray daily among U.S. states before and during the pandemic [6]. The focus on individual behavior and outcomes ignores the role of government in improving public health historically and the role of religion and science in the process [6].

There is a large variation in recent leading causes of death (cancer, heart disease, and COVID-19) among U.S. states. Data obtained for this study showed, in 2018-2019, the range of pooled age-adjusted rates among the states was: cancer (120-180), heart disease (120-248). In 2020-2021, the range for COVID-19 deaths was 10-fold (14-140). This paper reports a cross-sectional analysis of U.S. state differences in the mentioned age-adjusted death rates by cause before the pandemic (2018-2019) and COVID-19 during the first two years of the pandemic (2020-2021), correlated to prayer, politics, economic factors, physician accessibility, public health expenditures per capita, and urban residency. Infant mortality per 1000 live births was added as it has been correlated to inequality [23]. Although there is a huge variation in ideology, organization, and ceremonial attendance among religions, respondents to a US survey found 30-42 percent of Catholics, Protestants, and “other” religions pray for health [24]. The variations in the rates among 50 U.S. states are examined separately for each cause of death to test the hypothesis that prayer is associated with death risk, controlling statistically for the other factors. Correlations of daily prayer with health-related behaviors (smoking, diet, and COVID-19 vaccinations) were also examined using the same methodology.

## Materials and Methods

Eight hypothesized predictors of health effects were examined for their correlation to age-adjusted state death rates and behavioral risk factors – percent who pray once or more daily, Republican influence on state health policies as indicated by the percentage vote for Trump in 2016, percent households in poverty, median family income divided by a cost-of-living index, the Gini income inequality index, urban concentration of the population, physicians per capita, and public health expenditures per capita. The references to data sources of the mentioned variables are noted in Table 1.

**Table 1.** Data Sources.

|  |  |
| --- | --- |
| Daily Prayer | Lipka and Wormald, 2016, Pew Research Center [25] |
| Trump Vote 2016 | CNN, 2016 [26] |
| Median family<br>Income/cost of living | Moneyrates.com, 2019 [27] |
| Income Inequality<br>(Gini) | U.S. Census Bureau, 2019 [28] |
| Physicians per<br>population | National Center for Health Statistics, 2019 [29] |
| Percent households<br>with poverty income | U.S. Census Bureau, 2020 [30] |
| Percent Urban | U.S. Census Bureau, 2024 [31] |
| Public Health<br>Expenditures<br>Per Population | Shadac.org, 2024 [32] |
| Age-adjusted Mortality<br>Rates | National Institute of Minority Health and Health Disparities, 2018-2021 [33] |
| Infant Mortality Rate | National Center for Health Statistics, 2018-2019 [34] |
| Smoking, vegetable,<br>and fruit consumption | Centers for Disease Control Behavioral Risk Factor Surveillance System, 2024 [35] |

After examination of the correlations of potential predictor variables for collinearity of the mentioned predictor variables, least-squares regression estimates of their potential effects on death rates and behavioral risk factors were calculated. The form of the equation is:

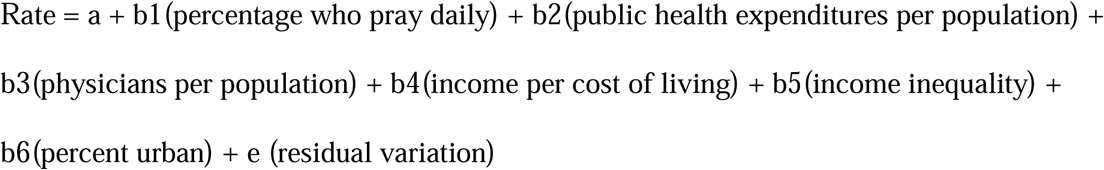

where “Rate” is a death rate or percent health behavior, as appropriate. In a separate analysis, the percentage of the vote captured by Trump in 2016 was substituted for the percentage who pray daily in the equations.

The Pew survey of religious behavior is based on a sample of about 35000 adults, about 700 per state, which produces a percentage for each state. The Behavioral Risk Factor survey is based on interviews with 2500 or more adults in each state. All of the data in this study were statistics downloaded from publicly available sources. There are no ethical risks to individuals.

## Results

The means and standard deviations of the variables included in the analysis are shown in Table 2. The standard deviations indicate substantial variation in each variable among the 50 U.S. states.

**Table 2.** Means and Standard Deviations of Analyzed Variables from 50 U.S. States.

| Variable | Mean | Standard<br>Deviation |
| --- | --- | --- |
| Percent Pray Daily | 54.2 | 8.8 |
| Percent Trump Vote 2016 | 49.7 | 10.2 |
| Median Income/cost of living | 906.6 | 101.5 |
| Inequality (Gini Index) x 100 | 46.4 | 1.8 |
| Percent Poverty | 12.1 | 2.7 |
| Physicians/100000 Population | 272.6 | 55.4 |
| Public Health<br>Expenditures/pop (\$) | 42.1 | 29.2 |
| Percent Urban | 72.4 | 14.8 |
| Cancer deaths/100000 population | 149.0 | 14.2 |
| Heart disease deaths/ 100000 population | 169.6 | 30.6 |
| COVID-19 deaths / 100000 population | 68.3 | 28.3 |
| Infant Mortality/ 1000 live births | 5.8 | 1.1 |
| Percent Who Smoke Cigarettes | 18.5 | 3.5 |
| Percent Consuming Recommended Fruit | 11.7 | 1.8 |
| Percent Consuming Recommended<br>Vegetables | 9.0 | 1.8 |
| Percent Fully Vaccinated Against<br>COVID-19 | 67.7 | 9.5 |

Correlations among the potential predictor variables were examined to avoid regression coefficients distorted by collinearity (Table 3). Most are not statistically significant. The most important are the correlations among physicians per population, the percentage who pray daily, poverty, and the Trump vote. On average, there are fewer physicians for potential access by the ill in states with more Trump voters and people who say they pray daily. The urban percentage is less correlated with the percentage of daily prayer than the Trump vote percentage. The percent who pray daily is strongly correlated to the percent in poverty, precluding their use as predictors in the same equation. While death rates are well known to be higher among the impoverished aged 40 and older [36], the percentage of the population who pray daily is three times that of the percentage in poverty (Table 2), encompassing a far larger population. The Gini inequality index is correlated to percent poverty, but not significantly to the other variables. It controls partly for poverty in the regression analysis.

**Table 3.** Least-squares correlation coefficients among predictor variables, N = 50 US states.

|  | Trump<br>Vote<br>2016 | Income/<br>Cost of<br>Living | Gini<br>Index | Percent in<br>Poverty | Physicians<br>/pop | Public<br>Health<br>Expend/Pop | Urban |
| --- | --- | --- | --- | --- | --- | --- | --- |
| Percent Pray<br>Daily | .59 | -.17<br>(ns) | .33<br>(ns) | .71 | -.57 | -.13 (ns) | -.13<br>(ns) |
| Percent Trump<br>Vote 2016 |  | .05<br>(ns) | -.12<br>(ns) | .45 | -.68 | -.25 (ns) | -.54 |
| Median<br>Income/cost of<br>living |  |  | -.23<br>(ns) | -.49 | .09 (ns) | -.36 | .17<br>(ns) |
| Inequality |  |  |  | .54 | .21 (ns) | -.08 (ns) | .26<br>(ns) |
| Percent Poverty |  |  |  |  | .31 | -.02 (ns) | .19<br>(ns) |
| Physicians/<br>Population |  |  |  |  |  | .10 (ns) | .25<br>(ns) |
| Public Health<br>Expenditures/<br>population |  |  |  |  |  |  | .04<br>(ns) |

ns = not statistically significant at p=.05, two-tailed.

Table 4 presents the regression coefficients of age-adjusted death rates per 100,000 population of the four selected causes of death with robust standard errors beneath the least-squares estimates. The left column for each cause of death contains the coefficients when prayer is included as a predictor, and the right column is when the percent Trump vote is substituted for the percent daily prayers. Contrary to the hypothesis that prayer reduces risk, consistently for each cause of death, more deaths per capita occur in states where people more often pray daily.

**Table 4.**
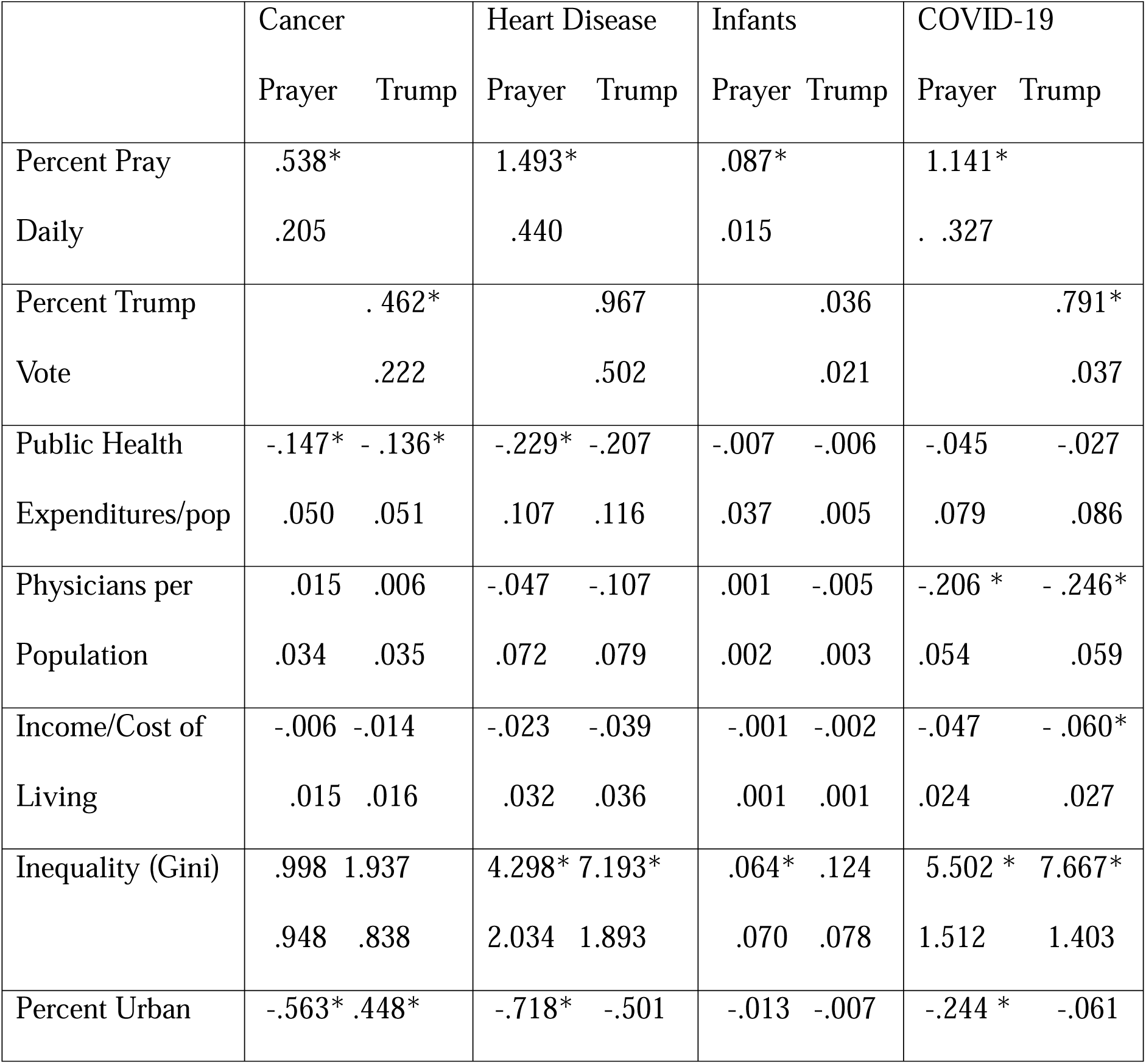
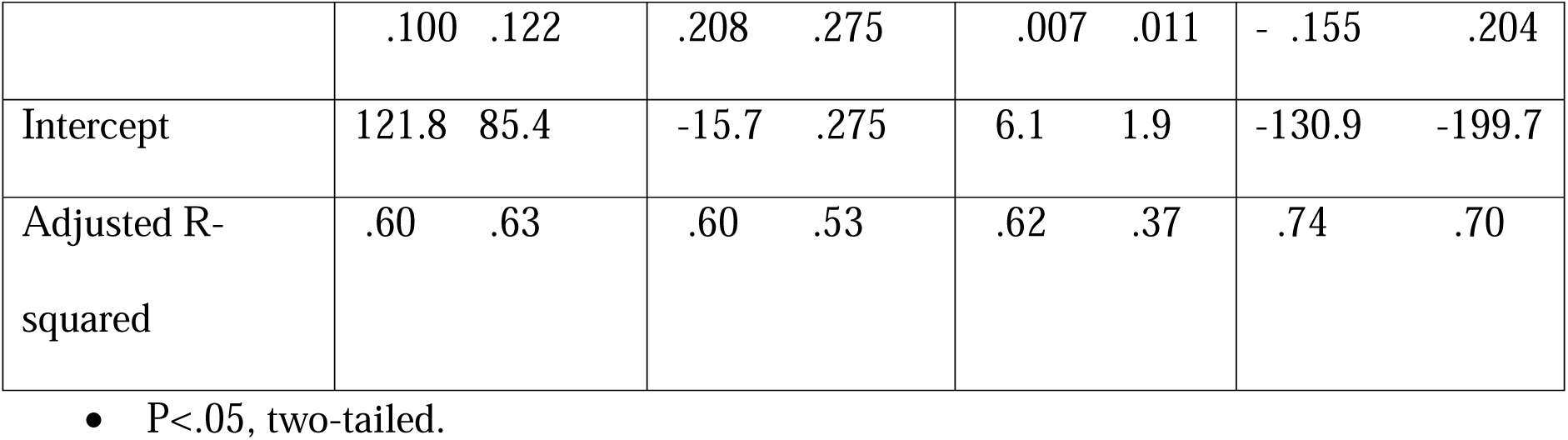
Least-squares regression coefficients and robust standard errors on predictor variables and selected age-adjusted mortality rates per 100,000 population, U.S. States 2018-2019, COVID-19 in 2020.

Prayer is the only variable significantly predictive of all four fatality rates. The Trump vote is not significantly associated with heart disease or infant mortality. Lower death rates from cancer and heart disease are associated with more public health expenditures, but not for infant mortality or COVID-19 deaths. COVID-19 death rates were lower in states with more physicians per capita, but that variable was not significantly associated with the other death rates. Heart disease, infant, and COVID-19 death rates were higher in states with more income inequality. All of the rates except infant mortality were lower in states where a greater percentage of the population lives in urban areas, but were only significantly predictive of cancer in the model that replaced the prayer variable with the percentage Trump vote.

The association of the predictor variables with important behavioral risk factors is displayed in Table 5. The greater the percentage of people who say they pray daily in a state, the more who report smoking cigarettes, consuming less than recommended fruits and vegetables, and having a lower percentage of the population vaccinated against COVID-19. Those behaviors were also reported by a greater percentage of people in states where Trump received a larger percentage of the vote and by a smaller percentage in states with more urban populations. Public health expenditures and physicians per capita were associated with a greater percentage of the population vaccinated against COVID-19, but not the other risk factors.

**Table 5.**
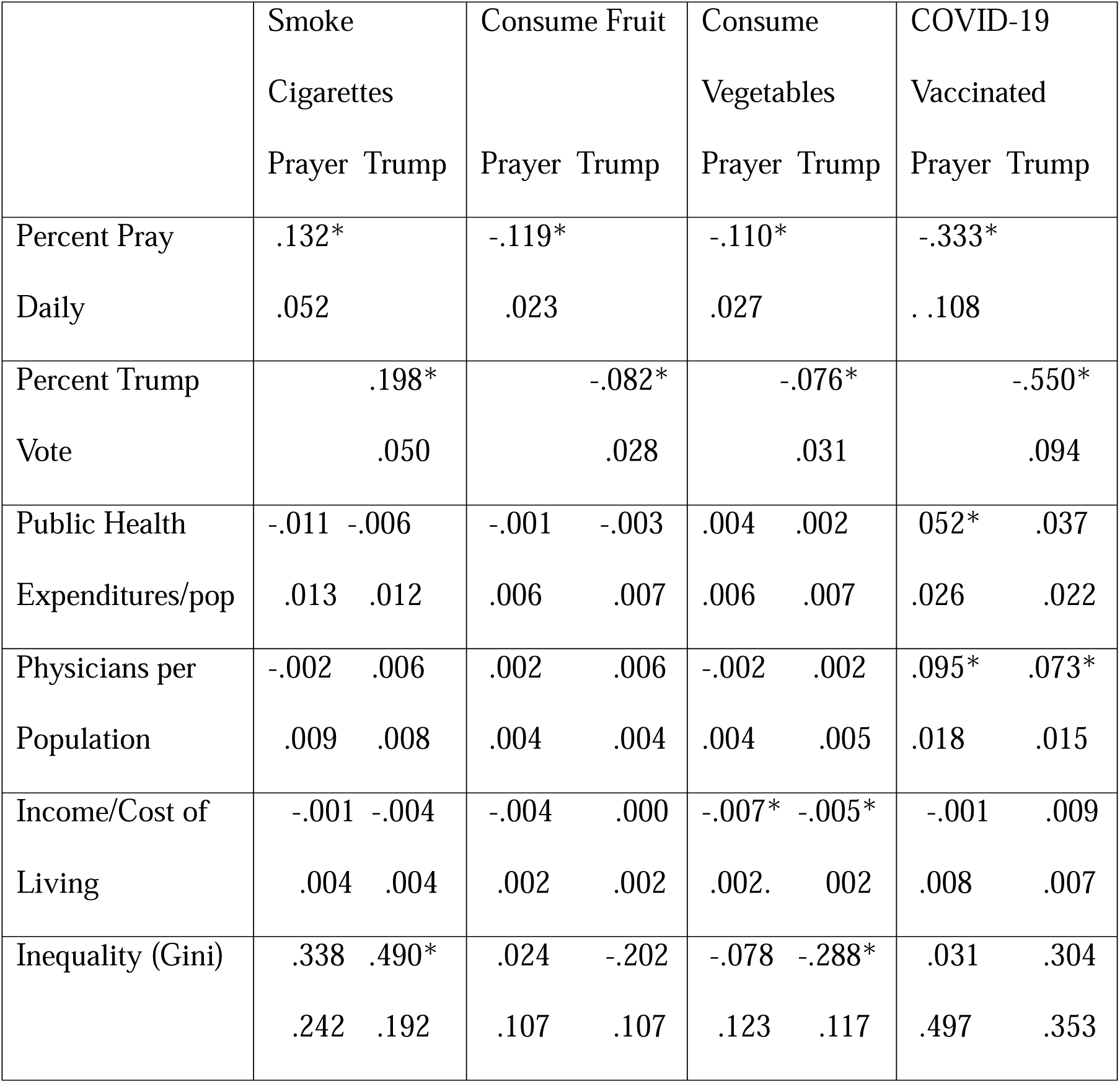
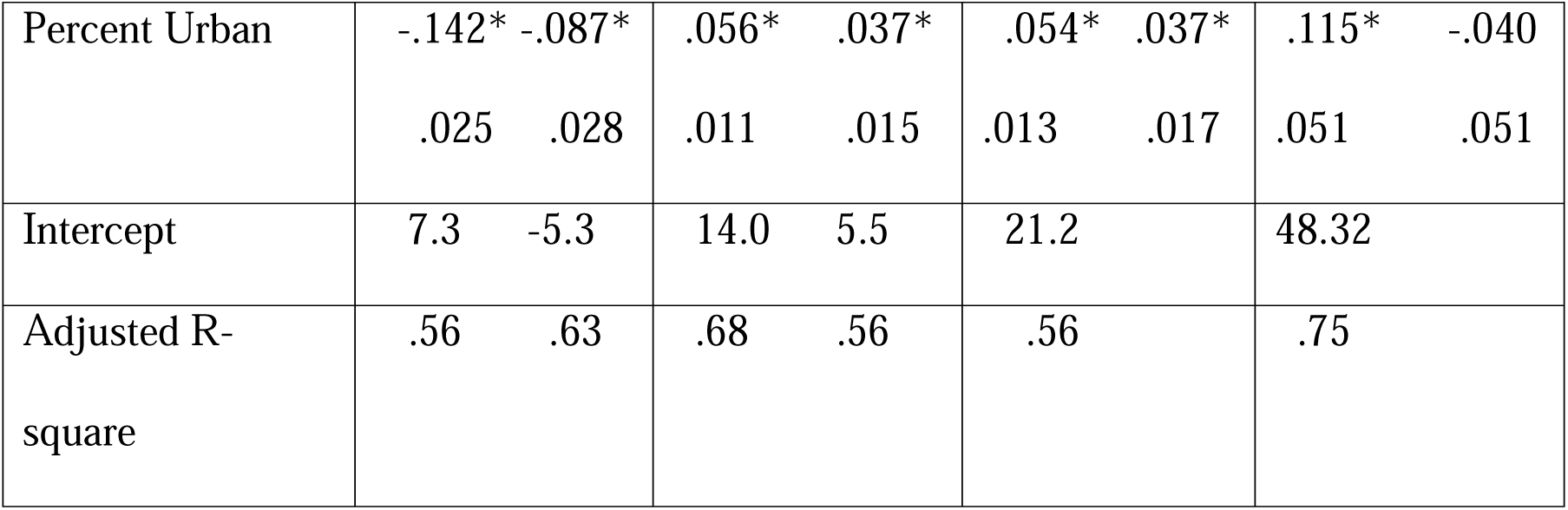
Least-squares regression coefficients and robust standard errors on predictor variables and the percent of selected health behaviors, N = 50 US states

## Discussion

The large variation in death rates among the states and their strong association with prayer frequency suggest that higher death rates could be at least partly because prayerful people are more likely to neglect preventive and ameliorative behavior. They may also be praying more due to poorer health. The correlation between daily prayer and cigarette smoking, as well as the inverse correlations between daily prayer and public health-recommended fruit and vegetable consumption, and COVID-19 vaccination, support the neglect hypothesis. The extent to which the neglect is an active rejection of public health recommendations or a lack of consideration of the evidence cannot be determined from aggregated data. As noted in the introduction, studies of health outcomes related to religious practices are mixed. The result on resistance to COVID-19 vaccinations is similar to that found in a review of twelve studies, finding that the religious are less likely to be vaccinated [37]. Thousands of lives were shortened because of vaccine resistance, and thousands more annually would be longer and healthier if there were less smoking and less consumption of foods that increase the risk of cancer and heart disease [38,39].

It is possible that the correlation between frequent prayer and death rates in this study partly results from prayerful people’s tendency to support state and federal governments that neglect less fortunate people and oppose environmental protection standards. For example, in a 2016 survey, the more prayerful were more likely to agree that aid to the poor “does more harm than good” and that “stricter environmental laws and regulations cost too many jobs and hurt the economy” [40]. Extensive research indicates that job loss related to environmental regulation is largely offset by jobs created to comply with and enforce the regulation [41].

Public health expenditures per capita are correlated with lower cancer and heart disease death rates, but not with reported health behaviors other than vaccinations. Since cancer and heart disease develop over extensive periods, current surveys of behavior may not reflect past changes, such as the large reduction in smoking [42]. In 2019, a state and local public health agency survey noted the percentage of particular programs. The most prominent were tobacco, alcohol, and drugs (74%), emergency preparedness (62%), infectious disease and vaccination (60%), food safety (48%), obesity/physical activity (45%), and wastewater/sanitation (39%). The agencies that served larger populations had higher percentages of each effort. State and local health departments can call on the Centers for Disease Control and Prevention when illnesses of unknown origin occur. Revenues to support public health departments are diversified – federal direct and passthrough (27%), local sources (25%), state sources (21%), Medicare/Medicaid (10%), and others combined (17%). These also vary by the size of the population served; state sources per capita are used more in less populated areas [43]. Therefore, it is not surprising that partisan politics is less correlated to public health expenditures than one might think. The anti-public health rhetoric and threats during the pandemic seem to have accelerated the trend of state and local public health professionals to retire or seek alternative careers. A study of changes in state and local public health staff trends from 2017 to 2021 does not portend well for the health of their states. Nearly half left the agencies, including three of every four younger than thirty-six [44].

During the first months of the newly elected Trump administration in 2025, Trump issued numerous executive orders that indiscriminately fired thousands of federal employees working in public health and other humanitarian efforts. Thousands of federally funded health-related research grants and contracts were canceled, continuing the anti-science trend in Republican politics [45]. The KFF Health Tracking Poll found that 72 percent of Republicans approved of the cuts [46].

Inferences from the lack of correlation between physicians per capita and death rates other than COVID-19 should be tempered by the possibility that the coefficients may be distorted by collinearity. Many medical prescriptions and procedures have been repeatedly verified for efficacy and safety in controlled clinical trials, but that does not guarantee that they are applied appropriately or that the recipients of treatment comply with instructions. Each must be judged based on evidence of correct application and compliance. The efficacy of medical care is partly offset by medical errors, deaths from which are estimated at more than 200,000 per year in the U.S. [47], competing with unintentional injuries as the third leading cause of death after heart disease and cancer.

The correlation of economic factors with death rates varies by mode of death. Corrected statistically for the other factors, income corrected for cost of living explains no variance. Income inequality is associated with each except for heart disease. Numerous studies have found higher death rates associated with poverty. COVID-19 deaths per capita were substantially higher among the poor and people engaged in essential low-wage work that could not be done remotely [48], but also people employed in health care [49].

A major limitation of this study is the inference of individual behavior from aggregate data. Occasionally, such inferences are found invalid. Also, there is the possibility that inclusion of one or more unmeasured variables would alter the results. While conclusive inferences of causation cannot be based on cross-sectional correlations, if prayer were efficacious in preventing deaths from the conditions examined, the correlations should be negative, not positive as they are. Of course, prayer does not cause the higher death rates in the more prayerful states, and increased prayer can be the consequence of chronic disease. An inference that a reduction in prayer, per se, would reduce disease would be false. The issue is the degree to which people neglect science-based prevention and treatment and support anti-science public policy because of religious beliefs. The correlations are of sufficient magnitude to warrant more detailed research on religious beliefs and practices regarding health maintenance, preventive strategies, treatment, and politics. Research on the effectiveness of eliciting clergy to promote improved health is also needed [50].

## Data Availability

All data produced are available online at the referenced sources.

